# Airway recommendations for perioperative patients during the COVID-19 pandemic: a scoping review

**DOI:** 10.1101/2021.07.30.21261372

**Authors:** Alexa Grudzinski, Billy Sun, MengQi Zhang, Agnes Crnic, Abdul H Djokhdem, Mary Hanna, Joshua Montroy, Laura V Duggan, Gavin M. Hamilton, Dean A Fergusson, Sylvain Boet, Daniel I McIsaac, Manoj M Lalu

## Abstract

**Purpose:** Numerous guideline recommendations for airway and perioperative management during the COVID-19 pandemic have been published. We identified, synthesized, and compared guidelines intended for anesthesiologists.

**Source:** Member society websites of the World Federation of Societies of Anesthesiologists and the European Society of Anesthesiologists were searched. Recommendations focused on perioperative airway management of patients with proven or potential COVID-19 disease were included. Accelerated screening was used; data extraction was performed by one reviewer and verified by a second. Data was organized into themes based on perioperative phase of care.

**Principal Findings:** Thirty unique sets of recommendations were identified. None reported methods for systematically searching or selecting evidence to be included. Four were updated following initial publication. For induction and airway management, most recommended minimizing personnel and having the most experienced anesthesiologist perform tracheal intubation. Significant congruence was observed amongst recommendations that discussed personal protective equipment. Of those that discussed tracheal intubation methods, most (96%) recommended video laryngoscopy, while discordance existed regarding use of flexible bronchoscopy. Intraoperatively, 23% suggested specific anesthesia techniques and most (63%) recommended a specific operating room for patients with COVID-19. Postoperatively, a minority discussed extubation procedures (33%), or care in the recovery room (40%). Non-technical considerations were discussed in 27% and psychological support for healthcare providers in 10%.

**Conclusion:** Recommendations for perioperative airway management of patients with COVID-19 overlap to a large extent. However, we also identified significant differences. This may reflect the absence of a coordinated response towards studying and establishing best-practices in perioperative patients with COVID-19.

**Registration:** Open Science Framework (https://osf.io/a2k4u/)

## Introduction

A defining feature of COVID-19 has been its rapid human-to-human transmission,^1^ largely through respiratory droplets and aerosols. Based mainly on retrospective evidence from the 2003 Severe Acute Respiratory Syndrome (SARS) epidemic, aerosol-generating medical procedures (AGMPs) such as tracheal intubation and extubation, and close proximity to the airway of perioperative patients were thought to increase the infection risk of anesthesiologists.^2^ In response, there has been a rapid proliferation of guidelines, recommendations, opinion pieces, checklists, and cognitive aids for the airway and perioperative management of patients with COVID-19. For the purposes of this review, we have labelled these diverse sources of information as ‘recommendations’. These recommendations have allowed perioperative health systems, anesthesiology societies, and departments of anesthesiology to establish policies and protocols to optimize the safety of both patients and healthcare providers.

The initial, rapid rise in recommendations may appropriately reflect regional differences in perioperative environments and resources. However, the publication of a large number of recommendations may limit knowledge translation, confuse providers, and potentially make it more difficult to adopt specific protocols. Contradictory information or discrepancies between recommendations may also decrease the perceived credibility of their sources, further delaying standardization of care. Indeed, the *choice overload hypothesis* would suggest that a larger number of choices may have negative motivating and affective consequences for behavior change.^3^

Given what we perceived to be a large number of guidance documents being published, we systematically identified and synthesized these perioperative airway management recommendations. This scoping review aimed to identify and review the breadth of recommendations for anesthesiologists and compare their content. Our scoping review also provides an important opportunity to assess one aspect of our community’s response to the COVID-19 pandemic and assist in establishing consistency while decreasing unnecessary practice variation as we prepare for the endemic phase of this disease.

## Methods

Our proposed scoping review followed a standard framework first defined by Arksey and O’Malley,^4^ expanded by Levac *et al*,^5^ and summarized by the Joanna Briggs Institute.^6^ Our protocol was posted *a priori* on Open Science Framework (https://osf.io/a2k4u/). Our final review is reported in accordance with the PRISMA extension for scoping reviews (PRISMA-ScR) (checklist in Appendix 1, Supplemental Digital Content).^7^

### Inclusion Criteria

To be eligible for inclusion in the review, sources had to meet the following criteria:

1. The target population included adult patients requiring surgery. Patient populations undergoing both cardiac and non-cardiac surgery were included.
2. Recommendations were for the perioperative airway management of patients with potential or proven COVID-19 disease. “Perioperative” was defined as any patient expected to have surgery, having surgery, or recently post-surgery, who are directly cared for by an anesthesiologist or anesthesia care provider.

### Exclusion Criteria

Sources that provided recommendations for perioperative airway management of patients with COVID-19 exclusively for obstetric, pediatric, or non-operative (e.g. endoscopy) populations were excluded. Sources that focused on airway management in non-operating room settings (intensive care units, trauma bay, hospital ward, prehospital care) were excluded.

### Literature Search

A search was conducted of national anesthesia organization webpages. We used the list of 136 member societies (representing 150 countries) of the World Federation of Societies of Anesthesiologists (WFSA) as well as the European Society of Anesthesiology and Intensive Care (ESAIC). Additionally, we searched the websites of other relevant organizations such as the Anesthesia Patient Safety Foundation and Safe Airway Society. Homepages of each society were opened in Google Chrome and, if needed, the translation function was activated. The homepage for each organization was reviewed for any information regarding COVID airway management guidelines, along with searches for “COVID” and “coronavirus” where webpage search functions were available. We extracted data from English, French, Croatian, Serbian, or Bosnian language material (languages spoken by the review team).

### Data Extraction

References were uploaded to DistillerSR (Evidence Partners, Ottawa, Canada), an audit-ready cloud-based program that assists with the design and conduct of systematic reviews. Full texts of included recommendations were screened independently and in duplicate. Initial disagreements were resolved by further discussion or, if necessary, a third senior author was consulted.

### Data Charting

The data abstraction form focussed on five general themes: PPE, airway equipment, non-technical skills, induction and extubation methods, and mental health of healthcare providers. Questions were refined via an iterative process involving all investigators and the data extraction form was then piloted using five of the included published recommendations (Appendix 2). Senior investigators reviewed the data abstraction of the five published recommendations to ensure the data abstracted was consistent with the research question and purpose as recommended by Levac *et al*.^*5*^ Data from included recommendations were transcribed into DistillerSR by one reviewer; a second reviewer independently verified accuracy of the data input. All data were verified by at least one team member with perioperative clinical experience.

To synthesize the results, data extracted was organized into five *a priori* determined themes based on perioperative phase of care: pre-operative, induction/airway management, intra-operative, extubation/recovery, and general recommendations (i.e. not specific to a phase of care). Data were exported to Excel (Microsoft, Redmond, USA). Frequencies of items are reported.

### Deviations from Protocol

In keeping with scoping review methodology, iterative changes were made in our protocol and have been documented in our registration. We originally searched MEDLINE, LitCovid, and Google for guidelines and recommendations surrounding the perioperative airway management of patients with COVID-19. These searches were conducted with the assistance of an information specialist (Risa Shorr, MLS, Ottawa Hospital Learning Services). As we updated our search, the number of captured publications proliferated. Number of records from our initial search increased exponentially from 94 in March 2020 to over 4000 by July 2020. In consultation with our information specialist, we modified our search to maintain feasibility resulting in 885 citations. Even with this modified search, 4951 records were retrieved by July 2021. Following screening, the majority of relevant publications identified were authored by small groups of individuals unassociated with national or international anesthesiology organizations. As such, our study group decided to restrict our search to websites of member societies of the WFSA and ESAIC to ensure that recommendations included in this scoping review were widely accessible and endorsed by national organizations or anesthesia societies, therefore potentially representing the highest quality and evidence-based recommendations as well as perceived as the most credible by clinicians for practice change. Furthermore, a single reviewer assessed whether included publications used systematic methods to search for evidence or described the criteria used for selecting evidence.

## Results

A total of 176 publications were retrieved from anesthesia society websites. Following screening, 30 publications, each representing a unique set of recommendations, met our eligibility criteria and were included in our analysis (Table 1).

**Table 1.** Study characteristics of included publications

| Society/Organization | Article Title | Country of Origin | Target Patient Population |
| --- | --- | --- | --- |
| Australian Society of Anesthetists | Anaesthesia and caring for patients during the COVID-19 outbreak | Australia | NS |
| Safe Airway Society | Consensus statement: Safe Airway Society principles of airway management and tracheal intubation specific to the COVID-19 adult patient group | Australia, New Zealand | Tested Suspected, Confirmed Positive |
| Safe Airway Society | COVID-19 Airway Management Infographics | Australia, New Zealand | Tested Suspected, Confirmed Positive |
| Society for Anesthesia and Resuscitation of Belgium | SARB recommendations for the anesthetic management of patients suspected of being infected or infected by the new coronavirus initially named 2019-nCoV | Belgium | Tested Suspected, Confirmed Positive |
| Association of Medical Practitioners Anesthesiologists-reanimatologists (UDMAR) | Coronavirus – guidance for anaesthesia and perioperative care providers | Bosnia and Herzegovina | Tested Suspected, Confirmed Positive |
| Canadian Anesthesiologists' Society | Canadian Anesthesiologists' Society COVID-19 Recommendations during Airway Manipulation | Canada | Tested Suspected, Confirmed Positive |
| Canadian Anesthesiologists' Society | Reinstitution of Elective Operations following COVID-19 | Canada | Untested Asymptomatic, Tested Suspected, Confirmed Positive |
| Chinese Society of Anesthesiology, Chinese Association of Anesthesiologists | Perioperative Management of Patients Infected with the Novel Coronavirus | China | Tested suspected, Confirmed positive |
| Chinese Society of Anesthesiology | Chinese Society of Anesthesiology Expert Consensus on Anesthetic Management of Cardiac Surgical Patients With | China | Untested Asymptomatic, Tested Suspected, Confirmed Positive |
|  | Suspected or Confirmed Coronavirus Disease 2019 |  |  |
| Société Française d'Anesthésie et de Réanimation | Proposals for the anesthetic management of a patient suspected or infected with Coronavirus COVID-19 | France | NS |
| Société Française d'Anesthésie et de Réanimation | Coronavirus 2019 et période périopératoire (COVID-19) | France | NS |
| Société Française d'Anesthésie et de Réanimation | Preconisations pour l'adaptation de l'offre de soins en anesthésie-reanimation dans le contexte de pandémie de covid-19-fiches-pratiques | France | Untested Asymptomatic, Tested Suspected, Confirmed Positive |
| Société Française d'Anesthésie et de Réanimation | Preconisations pour l'adaptation de l'offre de soins en anesthésie-reanimation dans le contexte de pandémie de covid-19 | France | NS |
| Italian Society of Anaesthesia, Analgesia, Resuscitation and Intensive Care (SIAARTI) | Surgery in COVID-19 patients: operational directives | Italy | Tested Suspected, Confirmed Positive |
| Kenya Society of Anesthesiologists | Kenya Society Of Anaesthesiologists Safe Anaesthesia Perioperative Preparation And Handling Of Confirmed Or Highly Suspected Covid-19 Patients (Protection in high risk aerosol generating procedures). | Kenya | Untested Asymptomatic, Tested Suspected, Confirmed Positive |
| Korean Society of Anesthesiologists | Management Of Critical Covid-19 Disease After Confirmation Of Tests | Korea | NS |
| Malaysian Society Of Anaesthesiologists & Malaysian Society Of Intensive Care | Recommendations For Management Of Anaesthesia And Intensive Care Services In Preparation Of Worsening Of The Covid 2019 Pandemic, | Malaysia | NS |
|  | Guidelines On Management Of Corona Virus Disease 2019 (Covid-19) In Surgery |  |  |
| Moroccan Society of Anesthesia and Resuscitation | Recommandations De Bonnes Pratiques Relatives A La Prise En Charge Des Cas Suspects Ou Confirmés Covid-19 | Morocco | Tested Suspected, Confirmed Positive |
| Serbian Association of Anesthesiologists and Intensivists | Coronavirus – guidance for anaesthesia and perioperative care providers | Serbia | Tested Suspected, Confirmed Positive |
| South African Society of Anaesthesiologists | Recommendations for the management of anaesthesia and surgery for Elective Procedures | South Africa | NS |
| South African Society of Anaesthesiologists | SASA Recommendations on Personal Protective Equipment (PPE) for anaesthesia providers during the COVID-19 pandemic | South Africa | NS |
| Taiwan Society of Anesthesiologists, Advanced Airway Committee | Endotracheal Intubation in Patients With COVID-19 Infection: Expert Panel-Based Consensus Recommendations | Taiwan | NS |
| Association of Anesthesiologists of Uganda | AAU/ICSU Covid-19 Airway management guidelines | Uganda | Untested Asymptomatic, Confirmed Positive |
| Association of Anaesthetists | Anaesthetic Management of Patients During a COVID-19 Outbreak | United Kingdom | Untested Asymptomatic, Tested Suspected, Confirmed Positive |
| Association of Anaesthetists, The Faculty of Intensive Care, Intensive Care Society, Royal College of Anaesthetists | COVID-19 airway management principles | United Kingdom | NS |
| American Society of Anesthesiologists | COVID-19 Information for Health Care Professionals | United States | Tested Suspected, Confirmed Positive |
| American Society of Anesthesiologists | COVID-19 Information for Health Care Professionals | United States | Tested Suspected, Confirmed Positive |
| Anesthesia Patient Safety Foundation, American | Perioperative Considerations for | United States | Untested |
| Society of Anesthesiologists, American Association of Nurse Anesthetists, American Academy of Anesthesiologist Assistants | the 2019 Novel Coronavirus (COVID-19) |  | Asymptomatic, Tested Suspected, Confirmed Positive |
| World Federation of Societies of Anaesthesiologists | World Federation Of Societies of Anaesthesiologists - Coronavirus | N/A | Tested Suspected, Confirmed Positive |
| World Federation of Societies of Anaesthesiologists | Perioperative management of suspected/ confirmed cases of COVID-19 | N/A | Tested Suspected, Confirmed Positive |

### Recommendation Characteristics

Original dates of publication were reported in 28/30 (93%), ranging from February 2020 to August 2020. We found updated versions for four publications (13%), with the most recent being updated in June 2021. Of these thirty, 23 (77%) referred to their publication as “recommendations”, and five (17%) referred to their publication as “guidelines”. None of the included publications used systematic methods to search for evidence or described the criteria used for selecting evidence. The majority (20/30, 67%) specified that the target patient population of their publication was confirmed COVID-positive patients, whereas 10/30 (33%) did not specify a target population (i.e., confirmed positive, suspected or asymptomatic, tested or untested).

### Phase of Care: Preoperative

A minority (7/30, 23%) of publications discussed the preoperative patient assessment. These included preoperative hygiene processes (e.g. hand hygiene) (7/30, 23%), and PPE for clinicians during preoperative assessment (4/30, 13%; Figure 2). Five publications (17%) recommended amendments to the usual pre-anesthetic history and physical examination; four suggested the addition of temperature measurement. Other recommendations included in these seven publications were a detailed risk assessment history (e.g., travel, COVID-19-specific symptoms and treatment), emphasis on chest examination and other organ systems affected by COVID-19 disease, and consideration of imaging such as chest x-ray or CT scan (especially in emergencies or if COVID status is indeterminate). One publication suggested the use of telemedicine when possible. Approximately half (14/30, 47%) of publications recommended PPE for patients, specifically the donning of a surgical/procedure mask.

**Figure 1.**
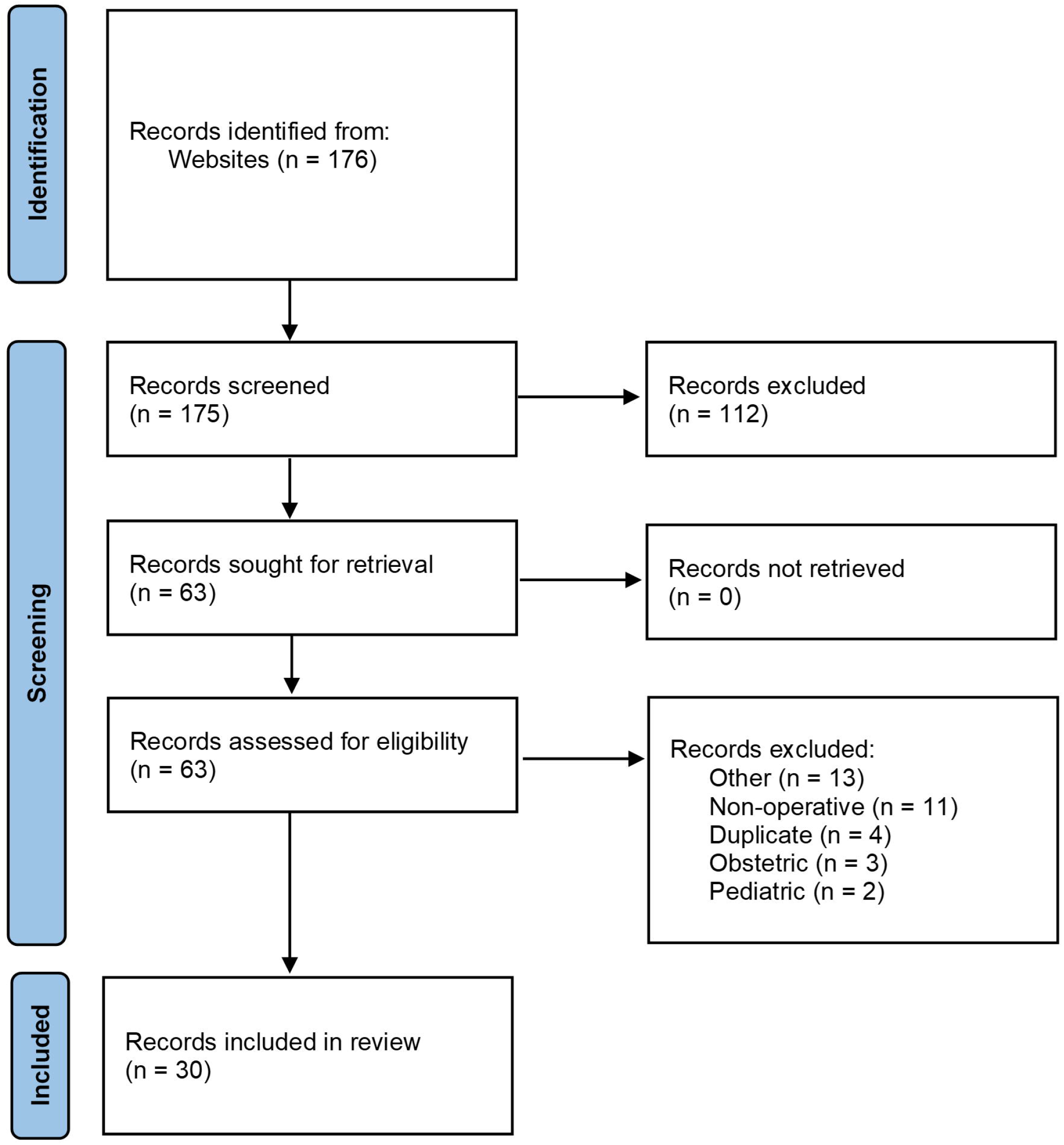
PRISMA flow diagram of recommendation selection.

**Figure 2.**
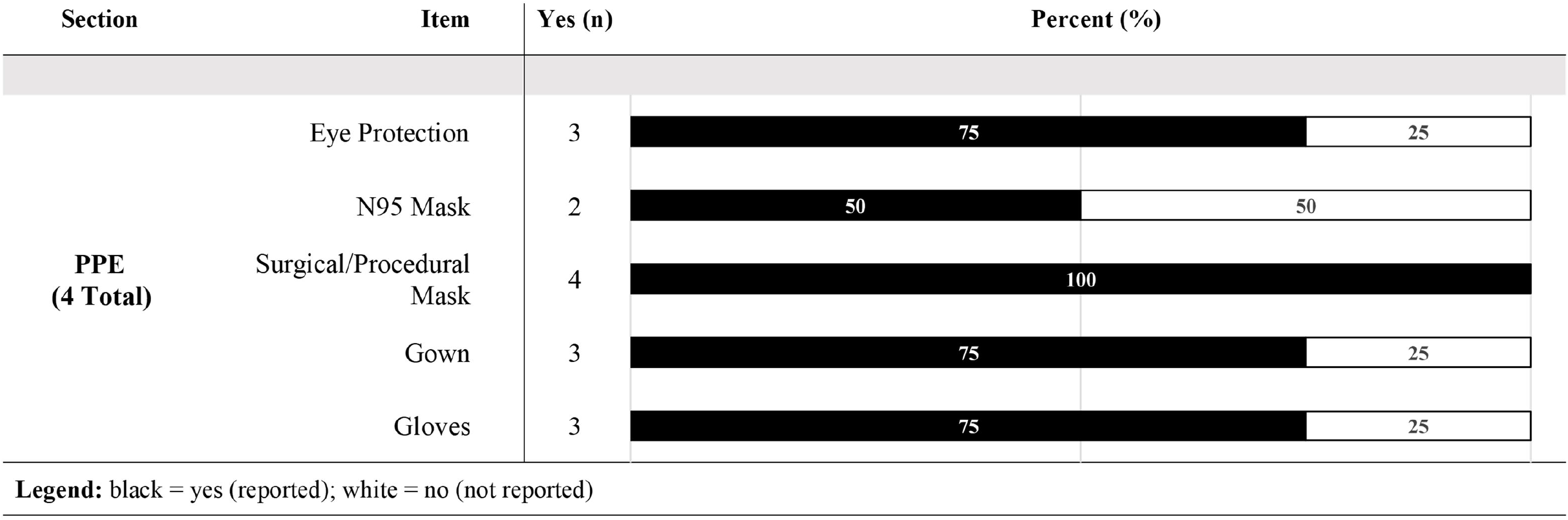
Summary of pre-operative assessment recommendations from the included publications (n=30).

### Phase of Care: Induction/Airway Management

The majority of publications (24/30, 80%) provided recommendations for specific team members and roles during anesthesia induction and airway management (Figure 3). Of these 24, 20 advised minimizing the number of personnel in the operating room to only those directly involved in establishing in the airway. Six publications provided specific suggestions for clearly defined roles (e.g., primary anesthesiologist, a second/assisting anesthesiologist, one “runner” within the OR, and one “runner” outside the operating room or in the ante-room). One article recommended a team leader distinct from the primary anesthesiologist to coordinate the team, manage drugs, observe monitoring, and provide airway assistance if necessary. The majority of publications 19/30 (63%) provided recommendations on who should perform tracheal intubation. All 24 publications recommended tracheal intubation be performed by the most experienced anesthesiologist. Of note, two of these publications provided recommendations to consider excluding staff who may be vulnerable (e.g., immunocompromised) from the airway team.

**Figure 3.**
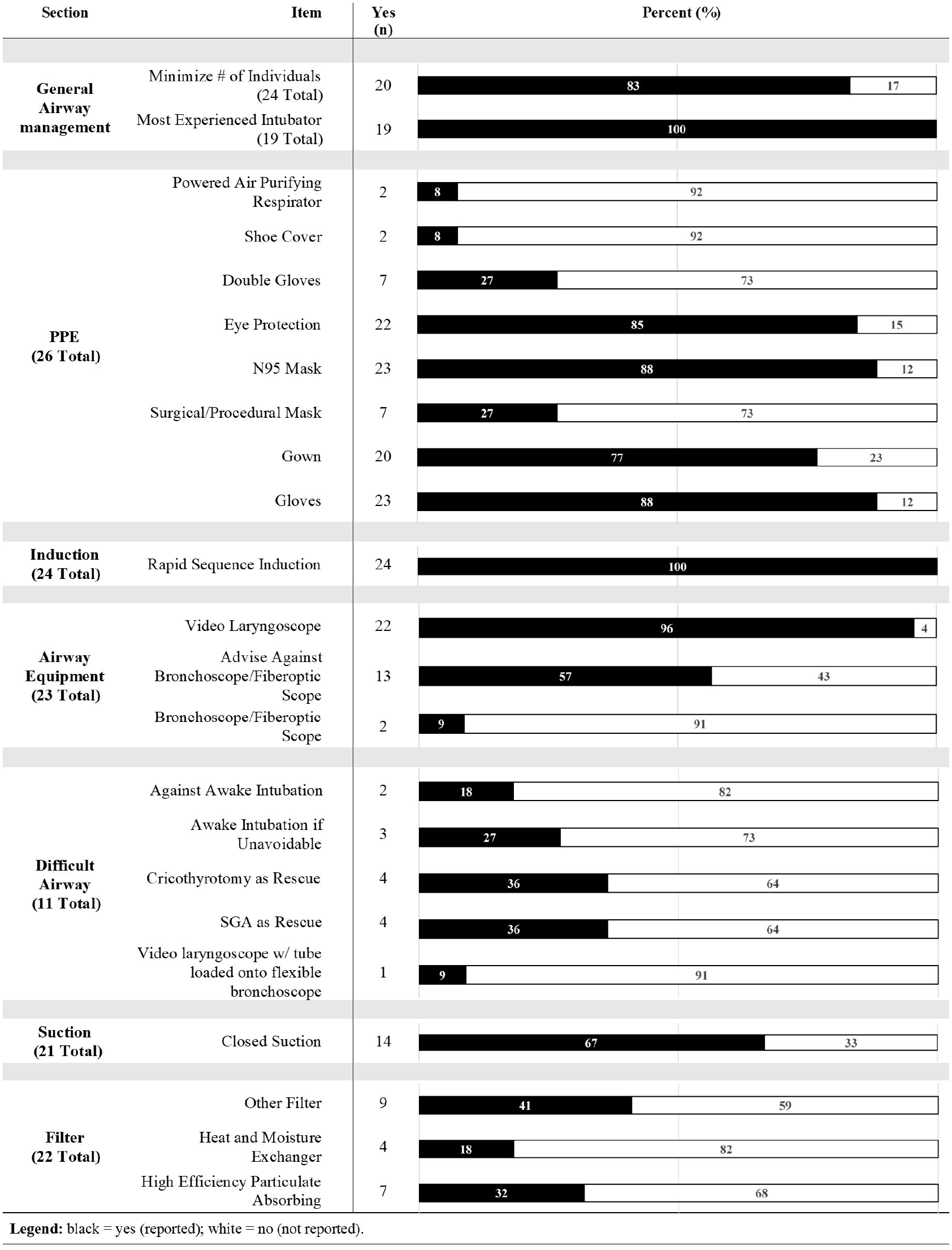
Summary of induction and airway instrumentation recommendations from the included publications (n=30).

#### PPE

The majority of publications (26/30, 87%) discussed PPE for anesthesia induction and airway management. PPE included gloves (23 publications), isolation gowns (20 publications), N95 masks (23 publications), and goggles or face shield (22 publications). Two publications discussed self-check for the donning/doffing of PPE in this phase of care, and six discussed PPE spotter-checks.

#### Airway Equipment

The majority of publications (23/30, 77%) provided specific suggestions for airway equipment, of which 22 (73%) recommended the use of video laryngoscopy. Thirteen publications specifically recommended against the use of flexible bronchoscopy for tracheal intubation; conversely, two publications supported its use. The majority of publications (22/30, 73%) provided specific suggestions surrounding use of a filter on the anesthetic circuit, with seven recommending a high efficiency particulate absorbing filter and four recommending a heat and moisture exchanger filter. Most publications (21/30, 70%) suggested the use of an intraoperative suction system, with 14 of these specifically describing a closed-suction system.

#### Preoxygenation

The majority of publications (20/30, 67%) discussed preoxygenation with all highlighting adequate preoxygenation specifically to avoid manual ventilation post-induction. Some suggested techniques to minimize potential aerosolization during preoxygenation included use of low flow oxygen (e.g. <6 L/min), two-handed facemask ventilation, a well-sealed/fitted facemask, and the application of wet gauze over the patient’s nose and mouth to block secretions.

#### Induction

Specific processes for the induction of anesthesia were mentioned in 24/30 (80%) publications, all of which recommended rapid sequence induction. Four publications specified dosing of neuromuscular blocking agents (rocuronium >1.2mg/kg total body weight or >1.5mg/kg ideal body weight, succinylcholine 1.5mg/kg total body weight). Two specifically recommended that sufficient time be provided for onset of neuromuscular blockade. Three publications discussed the use of pharmacologic adjuncts during induction of anesthesia to reduce the risk of aerosolization. Suggested agents included atropine or glycopyrrolate for secretion reduction and prophylactic anti-emetics.

#### Difficult Airway

Less than half of publications mentioned approaches to a difficult airway (11/30, 37%). With respect to awake tracheal intubation, two publications recommended against use of awake tracheal intubation techniques, while three stated awake techniques could be used if required. With respect to airway rescue techniques, four recommended the use of a supraglottic airway as a rescue device for hypoxia and four discussed the use of cricothyroidotomy in a cannot-intubate-cannot-oxygenate scenario.

### Phase of Care: Intraoperative

#### Type of Anesthesia

A minority of publications (7/30, 23%) recommended a specific type of anesthetic technique (Figure 4). Of these seven, four suggested the use of regional/neuraxial techniques when possible. Of note, most publications recommended the donning of a surgical/procedure mask on non-intubated patients during surgical procedures.

**Figure 4.**
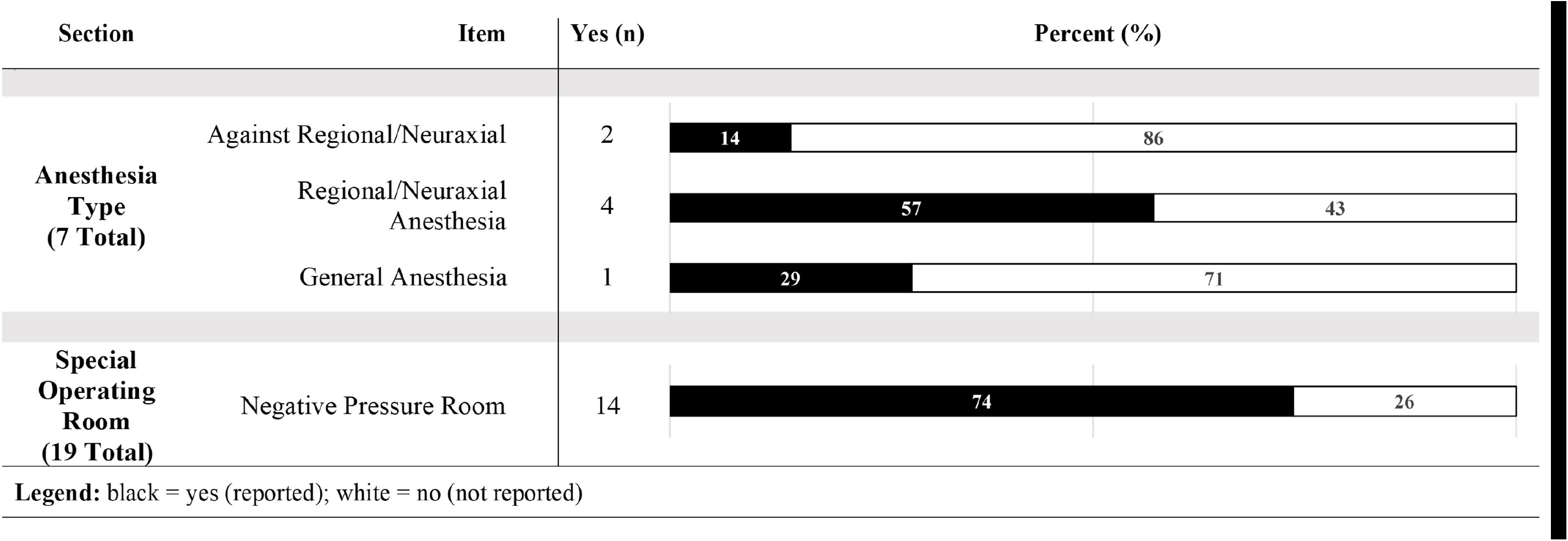
Summary of intraoperative management recommendations from the included publications (n=30).

#### Operating Rooms

Nineteen of thirty (63%) of publications mentioned having a specific operating room for patients with COVID-19. Of these publications, 14 recommended using a negative pressure operating room (Figure 4). In situations where a negative pressure operating room was not available, several publications recommended turning off the positive pressure and air conditioning systems commonly present in standard operating rooms. Labelling COVID operating rooms with specific signage was recommended in 16 publications. PPE for the intra-operative phase of case was discussed in two publications. Operating room disinfection was mentioned in 9/30 (30%) publications. Of these, three suggested implementing a waiting period between operations (e.g. 30-60 min). Two publications recommended a minimum rate of air exchange (12 and 25 exchanges per hour).

#### Equipment Cleaning

Nine publications (30%) commented on anesthesia machine cleaning. Of these publications, one recommended the use of disposable anesthetic machine covers and four recommended the use of “hospital approved” or EPA-approved disinfectant for anesthesia machine cleaning. Seven publications recommended that high-touch surfaces of the anesthetic machine be cleaned and disinfected and 20/30 (67%) recommended disinfection of equipment and/or the use of single-use equipment. Proper disposal of medical waste was mentioned in 14/30 (47%, e.g., a designated, labelled COVID-19 waste bin).

### Phase of Case: Postoperative

#### Extubation

Extubation was discussed in 10/30 (33%) publications, with recommendations for the specific extubation procedure provided in six (Figure 5). The most common suggestion for extubation involved the use of anti-tussive techniques. Specifically, one article recommended use of spontaneously breathing deep extubation and laryngeal mask airway exchange using a closed system.

**Figure 5.**
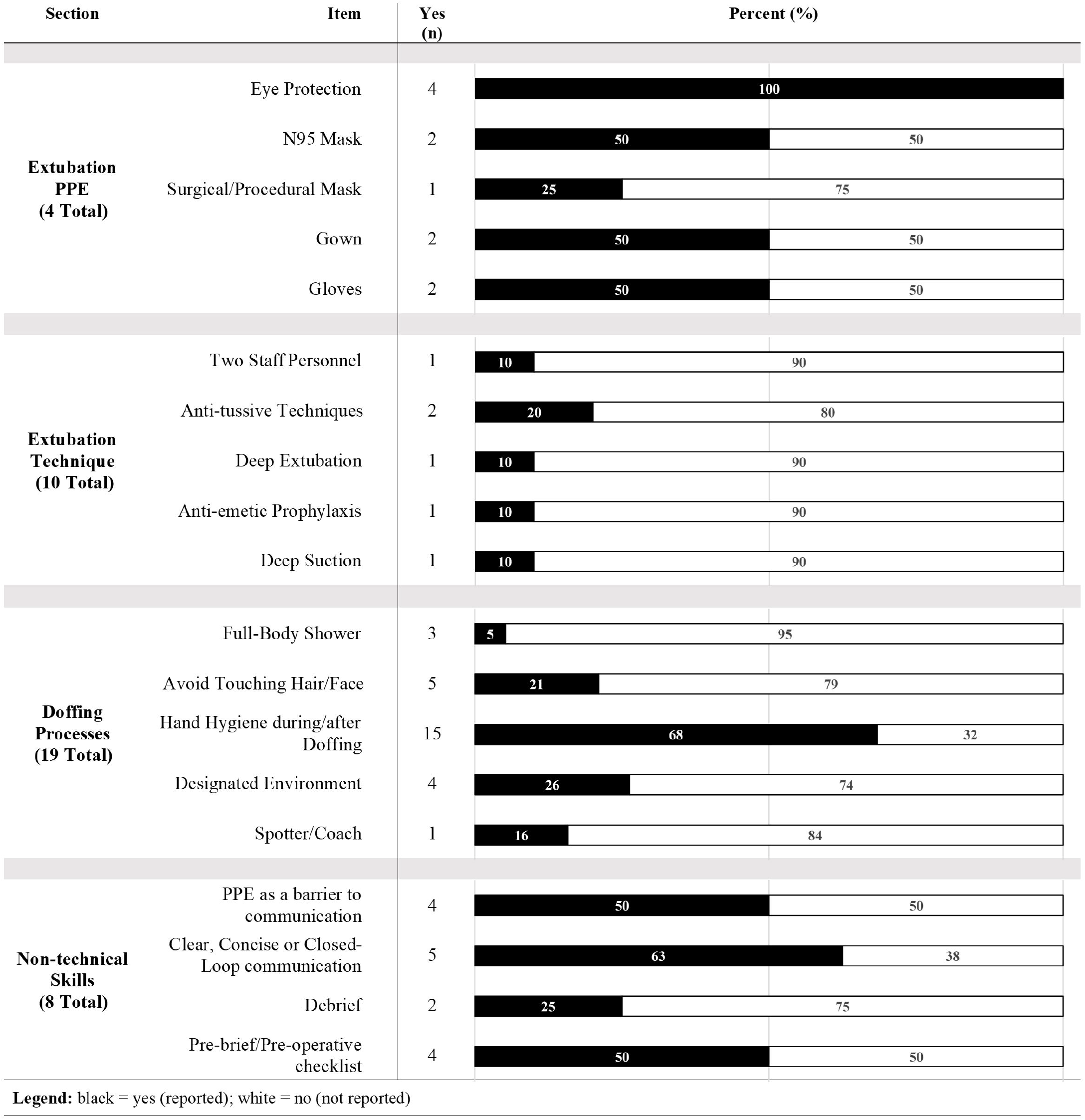
Summary of post-operative management recommendations from the included publications (n=30).

#### PPE

PPE for extubation was mentioned in 4/30 (13%) publications, the most common including the donning of gloves, an isolation gown, an N95 mask, and goggles or face shield (Figure 5). Doffing processes were discussed in 19/30 (61%) publications (Figure 5). Amongst these publications, 13 highlighted the importance of proper hand hygiene after doffing. Five publications recommended a designated environment for doffing, four highlighted the need to avoid self-contamination (i.e., touching their own face/hair) and three recommended the presence of a spotter/coach.

#### Post-anesthesia Care Unit

Fourteen publications (47%) discussed patient transport to recovery and twelve (40%) discussed care in the post-anesthesia care unit. General recommendations included wearing appropriate PPE, minimizing transport time, donning of a mask by the patient, and the use of a non-permeable patient cover during transport. Recommendations for tracheally intubated patients included minimizing the number and duration of breathing circuit disconnections, adequate paralysis, and clamping of the endotracheal tube during movement from one closed ventilation system to another to prevent aerosolization. One publication suggested a single dose of a 5-hydroxytryptamine receptor antagonist to prevent postoperative nausea and vomiting. A minority of publications discussed PPE during recovery room care (4/30, 13%) the most common of which was the use of gloves (2 publications), an isolation gown (2 publications), a surgical/procedure mask (2 publications), and eye protection (2 publications).

#### Support, Surveillance, and Non-Technical Considerations

Psychological support for healthcare providers (HCPs) was discussed in 3/30 (10%) publications. Suggestions included having available psychologic support services for HCPs, having adequate time off-work between shifts, and preparing for mental and physical fatigue. Surveillance of HCPs for potential COVID-19 symptoms and exposures was discussed in 5/30 (17%) of publications, of which two recommended HCPs keep a clinical logbook. Non-technical considerations were discussed by 8/30 (27%) publications (Figure 5). Of these, four recommended the use of a pre-brief/pre-operative checklist and two recommended a debrief. Five publications highlighted the importance of clear, concise, or closed-loop communication, and four identified the use of PPE as an important barrier to effective communication. Twenty (67%) publications recommended training. Of those that recommended training, the majority (15/20, 75%) recommended PPE training (donning, doffing). 8/20 (40%) recommended simulation-based training and 8/20 (40%) specifically recommended infection control training.

## Discussion

We synthesized peri-operative airway management recommendations during the COVID-19 pandemic. The majority of identified recommendations had a strong focus on the induction/airway and intra-operative phases of care. Despite the number and scope of publications reviewed, there was substantial agreement between societies on numerous aspects of COVID-19 perioperative airway management. Most recommendations included similar PPE recommendations, approaches to tracheal intubation including minimizing personnel present and suggested having the most experienced airway manager perform tracheal intubation. There was substantial congruence regarding specific tracheal intubation techniques, with most publications recommending a rapid sequence technique with video laryngoscopy. It is reassuring that the majority of societies provided congruent recommendations on multiple key components of perioperative airway management and care. Consistency in recommendations provided to anesthesiologists may have eased clinical decision-making and improved adherence to best safety practices in the COVID-19 era.

Although most publications provide similar suggestions, heterogeneity remains. As an example, while some publications specifically recommended against fibreoptic tracheal intubation in the difficult COVID-19 airway, others supported its use. Furthermore, there was substantial discordance in recommendations for both extubation techniques as well as PPE use during extubation. These conflicting recommendations may result in significant confusion for the individual practitioner, department or healthcare system when adopting best-practice protocols, particularly in the acute setting where not all perioperative factors are controlled. Additionally, there appears to be a lack of guidance surrounding appropriate PPE to be worn during the intra-operative phase of care, specifically during the period after tracheal intubation. Moreover, there is a lack of consensus regarding how long induction/airway management PPE should be worn during the operative case. Our findings highlight the absence of a unified consensus for perioperative airway management in patients with COVID-19.

Similar to a previous study, most of the recommendations identified in our review were not developed in a methodologically rigorous manner.^8^ For instance, no included recommendations used systematic methods to search for evidence nor did any clearly describe the criteria used for selecting evidence (two major components of evidence-based guideline development).^9^ This is likely a result of the relatively rapid nature in which these recommendations were developed and published, especially in the early days of the pandemic wherein our collective understanding of the virulence, routes or duration of transmission, and life cycle of the SARS-CoV-2 virus was limited. The lack of methodologic rigour in the development of the included recommendations may limit their successful uptake and implementation.^10^ However, when a new virus becomes the basis of a pandemic, recommendations prior to such knowledge being available, are at times required. The Campbell Inquiry, a federal government inquiry into the healthcare response to SARS in 2003 provides some principles for approaching a new public healthcare threat. Judge Campbell emphasized the use of the precautionary principle, which dictates that when there are two conflicting recommendations during an unclear healthcare threat, that the safer approach be followed.^11^ This results in frontline healthcare providers trusting that the recommendations made by national or society structures are to protect them. Once more evidence is gleaned, or other protections such as vaccination are widespread, recommendations should change to reflect the current situation.

Of great concern is the lack of updates to the majority of recommendations. As our knowledge and understanding of COVID-19 continues to grow, these documents have not kept pace. For instance, significant concerns have been raised surrounding aerosolization of COVID-19 during tracheal intubation; however, recent evidence has suggested that tracheal intubation in paralysed patients may not be aerosol-generating^12^, unlike the era of SARS when patients were oftentimes not paralysed during tracheal intubation.^2^ High-resolution environmental monitoring in ultraclean operating rooms has suggested that detectable aerosols during tracheal intubation might be 15-fold lower than with extubation, and detectable aerosols during extubation might be 35-fold lower than a volitional cough.^12^ As another example, recommendations generally suggested that negative pressure rooms may provide greater protection during aerosol-generating medical procedures performed in patients with COVID-19.^13–16^ However, negative pressure rooms have since been suggested to contribute to the increased the risk of developing pulmonary aspergillosis (which is commonly observed in patients with COVID-19).^17, 18^ Considering the rapidly emerging evidence, it is not surprising that outdated recommendations remain readily accessible. This undoubtedly contributes to ongoing confusion about, and lack of adherence to, the most up-to-date best practices. Interestingly, our original searches of MEDLINE, LitCovid, and Google identified a significant number of recommendations authored by small groups of individuals unassociated with national or international anesthesia societies. Given that these documents are unlikely to be developed and supported by anesthesia organizations or societies, the utility of continuing to publish such recommendations at this point in the pandemic is unclear. We would suggest that the time and resources expended on these recommendations may provide greater benefit if directed towards developing an international set of consensus-based guidelines, for both care providers and patients alike. We would also suggest a systematic international registry to curate access and management to pandemic-related guidelines. This will allow for outdated recommendations to be quickly identified.

Our study has limitations. Firstly, our results provide an overview of the current literature rather than answer a single, predefined question. Secondly, the recommendations included in our scoping review were not comprehensively assessed for methodologic quality (since this is not usually conducted for scoping reviews), although we would expect that included recommendations would likely score poorly in both methodological rigour and transparency of development.

Reflecting on our experiences with the COVID-19 pandemic, our study should inform future efforts to improve and streamline the pandemic response. A few central groups of critical care specialists were able to both create and organize emerging evidence and facilitate the creation of internationally applicable guidelines which could be locally modified as needed. For example, the REMAP-CAP and RECOVERY trials provided a global research platform to efficiently adapt to the COVID-19 pandemic, and rapidly evaluate various treatments in an expeditious manner.^19–21^ Similar international, multicentre initiatives within the perioperative research space were seldom observed, representing an important avenue of exploration to improve preparedness for future pandemics. Early synthesis and organization of evidence without redundancy in efforts will facilitate development of a judicious response to future pandemics. Our scoping review may be considered as one of the first needed steps to learn from experience. Just like individuals learn from reflexive practice,^22^ institutions are increasingly becoming learning organizations and we would like to suggest that specialty societies may adopt the same approach to become “learning societies” to optimize future patient care.^23^ By learning the available lessons during the COVID-19 pandemic, we may dramatically reduce the lead time to robust recommendations in healthcare while potentially avoiding the deleterious impact of pandemic-related restrictions. For example, the widespread cancellation of semi-urgent and elective surgeries in Ontario at the onset of the pandemic occurred in anticipation of a surge in patients with COVID-19.^24^ The complete impact of these increased wait times on population health has yet to be determined, and is ongoing.^25^ An important first step involves analyzing and identifying gaps in our current methods of creating and updating clinical practice guidelines, with a particular focus on how these processes should be refined in the context of a global pandemic and as COVID-19 evolves into an endemic disease.

## Supporting information

Supplemental Digital Content

## Data Availability

The datasets generated during and/or analysed during the current study are available from the corresponding author on reasonable request.

## Acknowledgements

The authors thank Risa Shorr, MLS (Learning Services, The Ottawa Hospital) for conducting the systematic searches.

